# A comparison study of SARS-CoV-2 IgG antibody between male and female COVID-19 patients: a possible reason underlying different outcome between gender

**DOI:** 10.1101/2020.03.26.20040709

**Authors:** Fanfan Zeng, Chan Dai, Pengcheng Cai, Jinbiao Wang, Lei Xu, Jianyu Li, Guoyun Hu, Lin Wang

**Affiliations:** Department of Clinical Laboratory, Union Hospital, Tongji Medical College, Huazhong University of Science and Technology, Wuhan, Hubei, PR China; Department of Immunology, School of Basic Medicine, Tongji Medical College, Huazhong University of Science and Technology, Wuhan, Hubei, PR China

## Abstract

**Objective:** To compare the difference of SARS-CoV-2 IgG antibody between male and female COVID-19 patients and figure out a possible explanation for different outcome between male and female patients.

**Methods:** A total number of 331 patients confirmed SARS-CoV-2 infection were enrolled. The plasma of these patients were collected during hospitalization and were detected for SARS-CoV-2 IgG antibody. Afterwards, the difference of IgG antibody between male and female patients was analyzed.

**Results:** The level of IgG antibody in mild, general and recovering patients showed on difference between male and female. In severe status, the average IgG antibody level in female patients tended to be higher than that of in male patients. Compared with male patients, most of the female patients generated a relatively high level of SARS-CoV-2 IgG antibody in severe status. In addition, the generation of IgG antibody in female tended to be stronger than male patients in disease early phase.

**Conclusions:** The inconsistent of SARS-CoV-2 IgG antibody generation in male and female patients may account for the different outcome of COVID-19 between gender.

## Introduction

In December 2019, an acute respiratory disease caused by an unknown respiratory syndrome coronavirus (SARS-CoV)-2 appeared in Wuhan, China[1]. The pneumonia has recently officially named as Corona Virus Disease 2019 (COVID-19) by World Health Organization (WHO). The incubation time of COVID-19 is generally 3 to 7 days, and the typically clinical manifestations are fever, cough, shortness of breath and fatigue.[2, 3]. By March 20, 2020, more than 80,000 confirmed cases and 3,000 dead cases have been reported in China. Most of the patients develop a mild or general symptoms and only a small part of people progresses to a severe or critical disease[2]. Previous study reported that male patients were susceptible to develop a more severe symptoms and had a higher mortality rate compared with female patients[2, 4, 5]. However, the factors underlying such a difference remains uncovered.

To date, the study about SARS-CoV-2 antibody are rare. The detection of serum IgM and IgG antibody had just been added to the New Coronavirus Pneumonia Prevention and Control Program (7th edition) to diagnosis SARS-CoV-2 infection. Because of no effective treatment for the patients in severe and critical status, the plasma donated by recovering patients containing high levels of SARS-CoV-2 IgG antibody are now used for transferring treatment in clinic in China. As expected, the plasma treatment brought significant attenuation in disease symptoms[6]. All of this indicated that anti-SARS-CoV-2 IgG antibody is a protective antibody. However, whether the IgG level exists a difference between male and female patients has not been reported.

In this study, by detecting and analyzing SARS-CoV-2 IgG antibody in recovering, mild, general and severe status patients, we identified that most of the female patients had a high level of IgG antibody relative to male patients in severe status. In addition, the production of IgG antibody seemed to be stronger in female patients in the early phase of COVID-19.

## Methods

### Patients

All the cases used in this study were derived from Wuhan Union Hospital, Tongji Medical College of Huazhong university of Science and Technology. The general information, clinical diagnosis and pathogenic diagnosis of patients were extracted from electronic medical records system. All of the enrolled patients were confirmed cases and classified by the New Coronavirus Pneumonia Prevention and Control Program (7th edition).

### Ethics approval

This study was approved by the Ethics Committee of Wuhan Union Hospital, Tongji Medical College, Huazhong University of Science and Technology. All people enrolled in the study signed informed consent forms.

### Anti-SARS-CoV-2 antibody detection

Serum sample were collected from healthy people and COVID-19 patients and used for SARS-CoV-2 antibody detection. The IgG antibody were detected with CLIA (chemiluminescent immunoassay) by iFlash 3000 (YHLO biotechnology co., LTD, Shenzhen, China). The IgG antibody detection kit were cat: C86095G (YHLO biotechnology co., LTD, Shenzhen, China). The cutoff value of IgG antibody is 10. All operations were in strict accordance with manufacturer’s instructions.

### Statistics

In this study, GraphPad 6.0 was applied for mapping and data statistical analysis. Mann-Whitney U test was used to compare plasma IgG antibody levels between two groups. P value less than 0.05 was considered statistically significant.

## Results

In this study, a total number of 331 in hospital patients were enrolled, among which male and female patients were 127 and 204 respectively. These COVID-19 patients contained three disease severity status (mild, general and severe) and some recovering patients. The number of this four were respectively 22, 87, 22 and 200. In mild group, male patient accounted for 36.4% and female patients accounted for 63.6%. The average age of male (M) and female (F) patients were 45.2 and 42.2 years old. In general group, male patients and female patients took up for 42.5% and 57.5% respectively. The mean age of male and female patients were 46.2 and 49.4. In the severe patients, the enrolled male and female cases were equal and the average age were 59.4 and 63.1. The recovering patients were in the late phase of the disease and were once diagnosed as mild or general. For recovering cases, male and female patients occupied respectively 35.5% and 64.5%, and the average age were nearly the same. As the same with previously study reported, elderly people enrolled in our study were more susceptible to severe status.

**Table 1.**
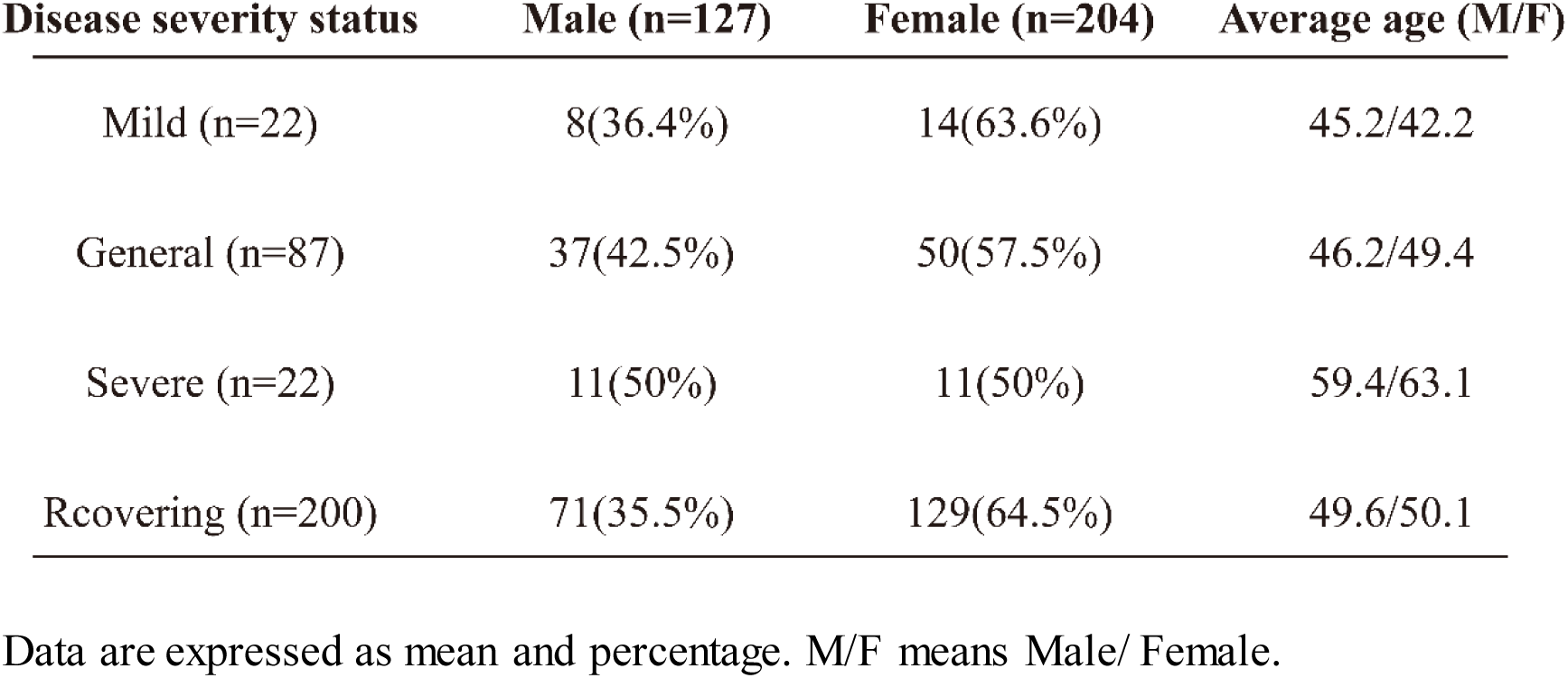
Basic information of patients enrolled.

| Disease severity status | Male (n=127) | Female (n=204) | Average age (M/F) |
| --- | --- | --- | --- |
| Mild (n=22) | 8(36.4%) | 14(63.6%) | 45.2/42.2 |
| General (n=87) | 37(42.5%) | 50(57.5%) | 46.2/49.4 |
| Severe (n=22) | 11(50%) | 11(50%) | 59.4/63.1 |
| Rcovering (n=200) | 71(35.5%) | 129(64.5%) | 49.6/50.1 |

Data are expressed as mean and percentage. M/F means Male/ Female.

To clarify the dynamic change of IgG antibody in male and female patients, we analyzed the level of IgG antibody between male and female patients in those four disease status. As shown in Figure 1A, in female patients, the level of IgG antibody continuously increased from mild status patients to general patients and severe patients, and then decreased in recovering patients. While in male patients, the IgG antibody raised from mild status patients to general patients, and then decreased from severe patients to recovering patients. In addition to severe status, the level of IgG antibody in the other three statuses were nearly the same. While in severe status, compared with male patients, the level of IgG antibody in severe status seemed to be higher in female patients. Next, we mapped scatter diagrams respectively for the above four status of patients. However, none of these four status showed a statistical difference in IgG antibody level between male and female patients (Figure 1B-1E). Notably, in severe status, most of the female patients had an antibody level for more than 100AU/mL, while in male patients, most of the IgG antibody were under 100AU/mL. Together, above data showed that the SARS-CoV-2 IgG level in female patients seemed to be higher than that of in male patients in severe status.

**Figure 1.**
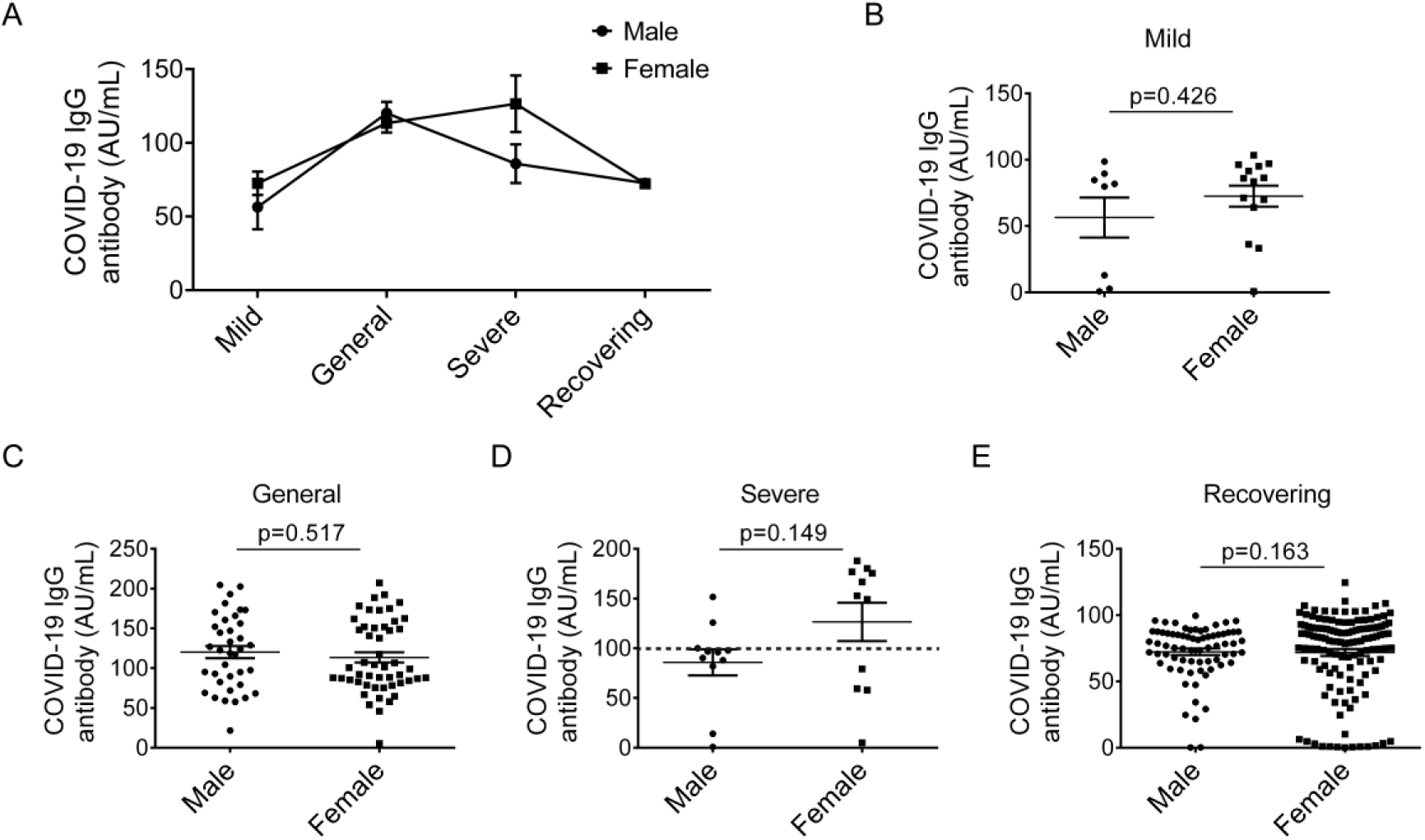
The level of COVID-19 IgG antibody in mild, general, severe and recovering COVID-19 patients. A) dynamic change of IgG levels in four status of disease of male and female patients. (B-E) comparison of SARS-CoV-2 IgG antibody between male and female patients in mild (B), general (C), severe (D) and recovering status patients (E). All data are expressed as mean ± SEM.

Next, to determine the concentration distribution of SARS-CoV-2 IgG antibody, we analyzed the proportion of the antibody in each range (0-10 AU/mL, 10-100 AU/mL, 100-150AU/mL and 150-200 AU/mL) in male patients and female patients. As is shown in Figure 2A, the proportion of range 0-10 AU/mL were consistent in male and female patients and the proportion of range 100-150 AU/mL was also close between male and female patients. However, the range 10-100AU/mL and 150-200 AU/mL showed great difference between two groups. In range 10-100 AU/mL, the male patients accounted for as much as 63.64%, while the female patient took up 27.27%. In range 150-200 AU/mL, the female patients accounted for as much as 54.55%, while the male patients occupied only 9.09%. Then we divided the severe status patient into two groups (high and low level: 0-100 AU/mL and 100-200 AU/mL) by antibody concentration. The statistical analysis showed that the level of IgG antibody in female patients was of no statistical difference between male and female patients in low level groups, while in high level groups female patients was significantly higher than male patients. These data suggest that there being more female patients maintained a high level of IgG antibody relative to male patients in severe status. In addition, we collected the time information of each cases and analyzed the difference in IgG level after disease onset. The result showed that the level of IgG antibody in female patients tended to be higher than male patients in 2-4 weeks after disease onset, and the difference in antibody disappeared after 4 weeks of disease onset. Above data indicated that the generation of IgG antibody differs between male and female patients after disease onset. IgG antibody in female patient generated a high level in most severe cases and a stronger production in disease early phase.

**Figure 2.**
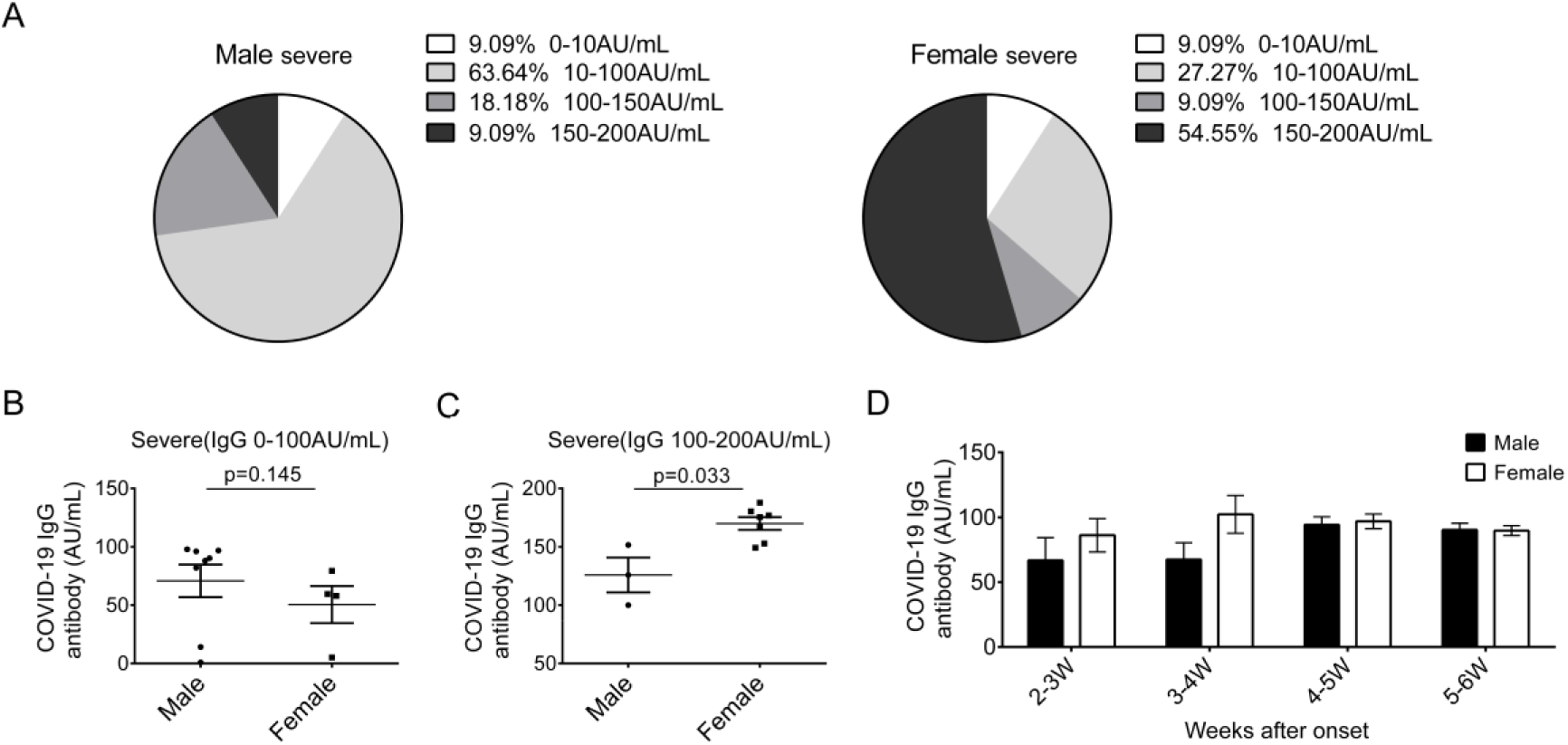
IgG antibody showed an accumulation in high levels in severe status and a strong production in early phase in female patients. (A) proportion of different IgG antibody concentration rage in male (left) and female (right) patients of severe status. (B-C) comparison and statistical analysis of IgG antibody divided into two groups, 1-100AU/mL(B) and 100-200AU/mL(C), in male and female patients of severe status. (D) the level of IgG antibody in indicate time points after disease onset. Patients were divided into four groups according to the weeks after disease onset. And the IgG level were analyzed in male and female patients. n (2-3W, 3-4W, 4-5W, 5-6W) =8, 10, 49, 42 for male and 11, 15, 48, 76 for female. All data are expressed as mean ± SEM.

## Discussion

Currently, the new coronavirus (COVID-19) pneumonia has widespread many countries in the world, caused more than 400,000 infections and 18,000 deaths. According to more than 7,000 cases that have been reported in China, a crude mortality of COVID-19 was about 2.3%. Among the diagnosed patients, about 80.9% were just showed mild to moderate symptoms and had a good prognosis[7]. However, for patients who had progressed to severe or critical status, the mortality rates significantly increased to as high as 49%[5]. And so far there was no effective treatment for COVID-19 patients, especially for severe and critical patients[8]. So it is urgent to distinguish and interfere in advance to prevent those patients who were likely to progress to severe and critical status.

Previous study reported a high severe rate and death rate in male patients relative to female patients[2]. However, the possible reason underlying was not referred. Here we reported that the production and dynamic of IgG antibody were different between male and female patients. In the early phase of the disease, the IgG antibody presented a stronger production in female patients. In the severe status, contrast with the male, SARS-CoV-2 IgG antibody in female tended to enrich in the high levels. The above two phenomena may be good reasons to explain the different outcomes between male and female COVID-19 patients. However, the exact mechanism contributing to this difference remains elusive and needs further investigation. At present, convalescent plasma was considered as a potential therapy for COVID-19 and now has been used in sever and critical patients in clinical[9]. Our study proposed that convalescent plasma should be used in advance especially for female patient with low IgG antibody for a long time to prevent the disease from progressing to severe and critical status, if the convalescent plasma is available. In addition, our study also supported that monitoring the SARS-CoV-2 antibody for routine examination maybe an efficient way to predict the progression of COVID-19 patients.

It should be noted that there were some limitations of this study. Firstly, as all the cases are collected in a signal hospital (Wuhan union hospital), the sample size was relatively small. Secondly, there were no critical cases enrolled in this study. Thirdly, probably due to limited sample size, the rate of severe cases for male and female patients did not showed a consistent tendency as previously reported. Nevertheless, the data showed in our study revealed a different pattern of IgG antibody after infection.

In conclusion, in this study we detected and analyzed the SARS-CoV-2 IgG antibody of 331 patients and found that, compared with male patients, more female patients maintained a high level of IgG antibody after SARS-CoV-2 infection. And the production of IgG in female patients tended to be stronger than male patients in early phase of COVID-19. We propose here that our doctors and nurses should pay more attention to the patients whose IgG antibody were at low levels. And monitoring IgG antibody maybe a potential method to assess COVID-19 progression.

## Data Availability

The data are available from the author upon request.

## Conflict of interests

The authors declare no competing interests.

## Acknowledgements

We thank all of the doctors, nurses and public health workers for their efforts in fighting against SARS-CoV-2 and saving the lives of COVID-19 patients. The project was supported by the Department of Clinical Laboratory, Union Hospital, Tongji Medical College, Huazhong University of Science and Technology.

## Contributions

Fanfan Zeng, Pengcheng Cai, Biaojin Wang, Lei Xu, Jianyu Li and Guoyun Hu contributed to the collection and detection of clinical samples. Fanfan Zeng and Chan Dai analyzed the data and wrote the manuscript. Lin Wang conceived and supervised the project

## Notes

### Competing Interest Statement

The authors have declared no competing interest.

### Funding Statement

There was no funding support in this work.

